# Effectiveness of Intravenous Immunoglobulin for Children with Severe COVID-19: A Rapid Review

**DOI:** 10.1101/2020.04.17.20064444

**Authors:** Jingyi Zhang, Yinmei Yang, Nan Yang, Yanfang Ma, Qi Zhou, Weiguo Li, Xia Wang, Liping Huang, Xufei Luo, Toshio Fukuoka, Hyeong Sik Ahn, Myeong Soo Lee, Zhengxiu Luo, Yaolong Chen, Enmei Liu, Kehu Yang, Zhou Fu, on behalf of COVID-19 evidence and recommendations working group

## Abstract

**Background:** Intravenous immunoglobulin (IVIG) is usually used as supportive therapy, but the treatment of COVID-19 by IVIG is controversial. This rapid review aims to explore the clinical effectiveness and safety of IVIG in the treatment of children with severe COVID-19.

**Methods:** We systematically searched the literature on the use of IVIG in patients with COVID-19, Severe Acute Respiratory Syndrome (SARS) or Middle East Respiratory Syndrome (MERS), including both adults and children. We assessed the risk of bias and quality of evidence and reported the main findings descriptively.

**Results:** A total of 1519 articles were identified by initial literature search, and finally six studies, included one randomized controlled trial (RCT), four case series and one case report involving 198 patients. One case series showed the survival of COVID-19 patients with acute respiratory distress syndrome (ARDS) was not improved by IVIG. One case report showed high-dose IVIG could improve the outcome of COVID-19 adults. Three observational studies showed inconsistent results of the effect of IVIG on SARS patients. One RCT showed that IVIG did not reduce mortality or the incidence of nosocomial infection in adults with severe SARS. The quality of evidence was between low and very low.

**Conclusions:** The existing evidence is insufficient to support the efficacy or safety of IVIG in the treatment of COVID-19.

## Background

Coronavirus disease 2019 (COVID-19), first reported in China on December, 2019, is an infectious disease caused by severe acute respiratory syndrome coronavirus 2 (SARS-CoV-2) (1). SARS-CoV-2 belongs to the family of coronaviruses, which are enveloped viruses that can cause illnesses ranging from common cold to severe diseases such as SARS and MERS (2,3). The COVID-19 epidemic massively influences the public health and people’s daily lives, and the disease was declared a pandemic on 11 March 2020 (4). All populations are susceptible to infection and there is a research shows children are as likely to be infected as adults (5). On February 11, 2020, there were 44672 confirmed cases in mainland China, of whom 416 were under the age of 10 years and 549 between the ages of 10 to 19 years (6). The main symptoms in children are fever and cough, and the disease is on average less severe in children than adults (7). However, severe cases have been reported also in children (8). So far, there has been no specific treatment for COVID-19, antiviral therapy and vaccination are currently under development (9,10).

IVIG is prepared from the plasma of healthy humans and usually used as supportive therapy. Its main component is immunoglobulin (Ig) G, which has dual therapeutic effects of immune-modulation effects and immune substitution (11). IVIG is one of the alternative treatments for children with agammaglobulinemia, and an effective treatment of Kawasaki disease (12,13). IVIG was used to treat SARS patients during the SARS outbreak in 2003 (14,15), but there is no convincing evidence of its effectiveness. According to recent reports, about 33% of patients with severe COVID-19 received IVIG in China (16). Some published guidelines of COVID-19 have indicated that IVIG could be used to treat children with severe or critical disease (17).

The purpose of this study is to perform a comprehensive rapid review to explore whether it is beneficial to treat children with severe COVID-19 with IVIG and provide supporting evidence support for COVID-19 guidelines. Because of the urgent situation, the review was not registered (18).

## Methods

### Search strategy

We carried out a comprehensive search in the following electronic databases: the Cochrane library, MEDLINE (via PubMed), EMBASE, Web of Science, China Biology Medicine disc (CBM), China National Knowledge Infrastructure (CNKI), and Wanfang Data, by using the terms “COVID-19”, “SARS-CoV-2”, “Novel coronavirus”, “2019-novel coronavirus”, “2019-nCoV”, “SARS”, “MERS”, “IVIG”, “intravenous immunoglobulin” and their derivatives. The search covered the time from each database’s inception to March 31, 2020. The search strategies are determined by multiple pre-searches and will be discussed with the clinicians about the appellation of disease and IVIG. We also searched the World Health Organization Clinical Trials Registry Platform, ISRCTN Registry, ClinicalTrials, Google Scholar, three preprint services, including medRxiv (https://www.medrxiv.org/), bioRxiv (https://www.biorxiv.org/) and SSRN (https://www.ssrn.com/index.cfm/en/) and references of included studies. The details of the search strategy can be found in the ***Supplementary Material 1***.

### Inclusion and exclusion criteria

We included RCTs that compared IVIG treatment (standard intravenous immunoglobulin preparations, excluding IgM-enriched immunoglobulin, hyperimmune immunoglobulin and specific immunoglobulin from convalecent plasma) with a control group (placebo or no treatment with IVIG), and cohort studies, cross-sectional studies, case-control studies, case series and cases report that can distinguish the corresponding outcomes caused by IVIG. The inclusion of COVID-19 adult patients and patients with SARS or MERS helps to provide indirect evidence, if studies on COVID-19 in children are scarce. Studies with all patients diagnosed with COVID-19, SARS or MERS were included, without restrictions on age, race, gender, or geographical location or setting. The primary outcomes were the risk of death and severity of the disease. Secondary outcomes included the incidence of nosocomial infection, the duration of hospitalization, clinical symptoms, absorption of lung lesions, improvement in abnormal biochemical indicators, and adverse effects. We excluded duplicates, conference abstracts, comments and letters, studies published in languages other than English or Chinese, and studies where we could not access the full text.

### Study selection

After eliminating duplicates by EndNote software and manual check, two reviewers (J Zhang and Y Yang) independently reviewed the titles and abstracts of records retrieved from the search and selected all potentially relevant studies according to the pre-defined inclusion and exclusion criteria. After this, the same reviewers screened the full texts and made the final selection. A pilot search was conducted before the full screening of the literature to ensure that each researcher understood the screening criteria and process. Disagreements about selection of studies were resolved by consulting a third party (N Yang). The process of study selection was documented using a PRISMA flow diagram (19).

### Data extraction

Two reviewers (J Zhang and Y Yang) independently extracted the following data from included trials using a standardized extraction sheet: 1) basic information (year of publication, first author and affiliation, journal, funding, conflict of interest); 2) study details (type of study, sample size, research purpose, research population characteristics, interventions; and 3) outcome data. A pre-test was conducted before formal extraction to ensure that each researcher agreed with the extraction criteria and process. Disagreements were solved through discussion with a third researcher (N Yang).

### Risk of bias assessment

Two reviewers (J Zhang and Y Ma) assessed the quality of the included studies independently. We used the Cochrane bias risk assessment tool (Risk of bias) to assess the randomized controlled trials and clinical controlled trials (20), the criteria recommended by the National Institute of Health and Clinical Optimization (NICE) for case series to assess the risk of bias (21), the Joanna Briggs Institute’(JBI) case report quality appraisal tool for case reports(22) Newcastle-Ottawa Quality Assessment Scale (NOS) for the quality of cohort studies and case-control studies (23), and Agency for Healthcare Research and Quality (AHRQ) tool for cross-sectional studies (24).

### Data synthesis

For dichotomous outcomes we calculated the risk ratio (RR) and the corresponding 95% confidence interval (CI) and *P* value. For continuous outcomes, we calculated the mean difference (MD) and its corresponding 95% CI when means and standard deviations (SD) were reported. If sufficient data were available, we considered examined the robustness of meta‐analyses in a sensitivity analysis.When effect sizes could not be pooled, we reported the study effectives narratively.

### Quality of the evidence assessment

The quality of the body of evidence was graded using the GRADE method (25,26). Evidence from randomized trials could be downgraded by the following five factors: risk of bias, inconsistency of results, indirectness of evidence, imprecision of results, and publication bias. The quality of evidence for each outcome was graded as high, medium, low, or very low. The results of the grading were presented in “GRADE evidence profile” (27-30).

## Results

We identified 1519 articles in the initial literature search (*Figure 1*). After removing duplicates, we screened the titles and abstracts of 1405 records. Thirty-one articles were retrieved for full-text reviewing. Finally, one RCT, four case series and one case report involving a total of 198 patients were included for rapid review (31-36).

**Figure 1.**
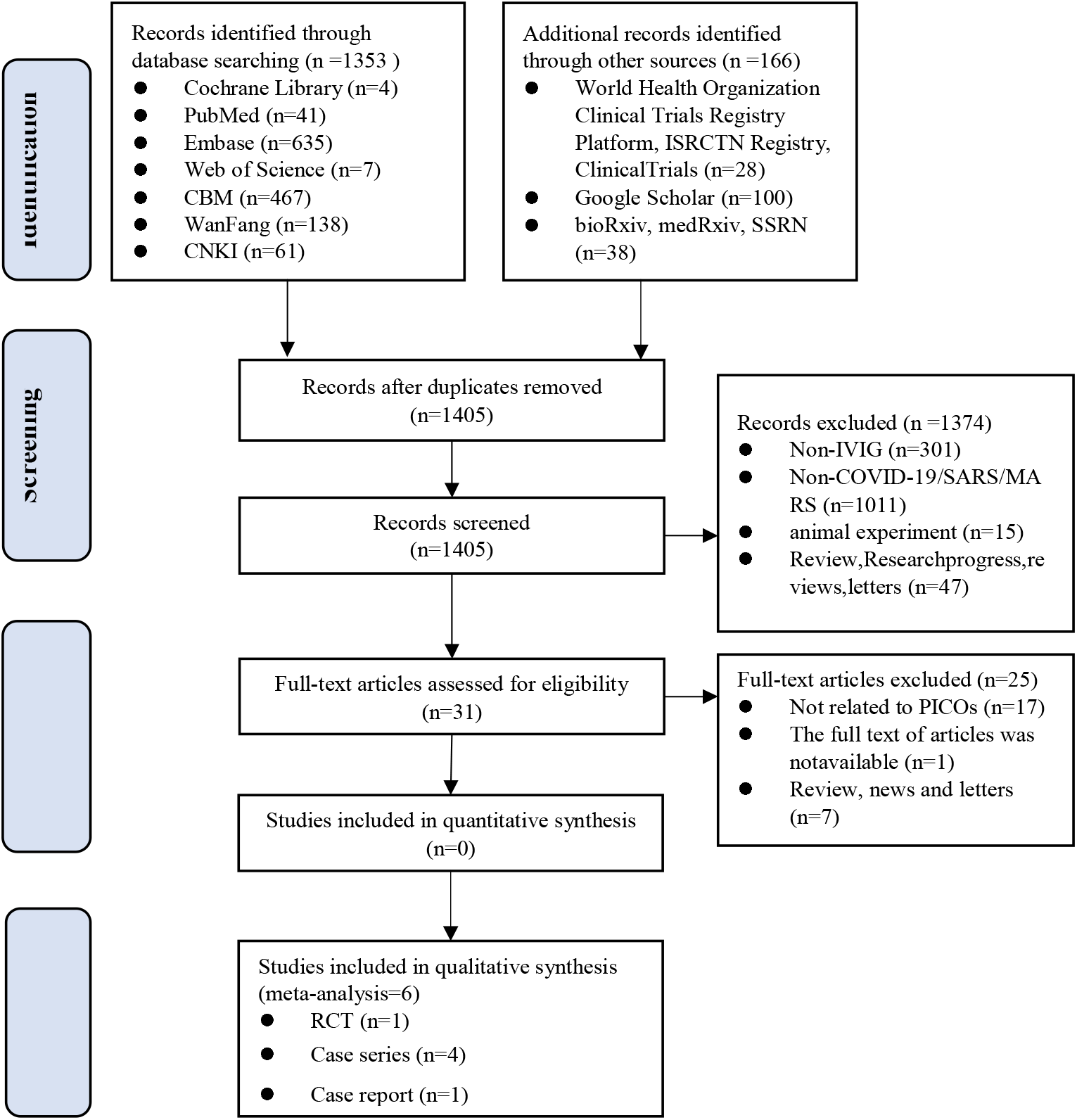
The flow chart of the literature search.

The studies were published between 2004 and 2020, and all studies were from China (*Table 1 and Table 3*). We found one case series on IVIG in COVID-19 adults with ARDS, one case report in COVID-19 adults, one randomized controlled trial of 44 adults with severe SARS, which included 25 adults with acute lung injury (ALI) and 19 adults with ARDS), one case series involving children with SARS, and two case series also involving adults with SARS. The outcomes of the studies were the duration of fever, total peripheral blood WBC, time of the lung lesions subsided obviously, adverse effects, WBC counts, platelet counts, serum globulin, the incidence of nosocomial infection, the risk of death, survival probability and the progression of disease cascade. In all studies IVIG was used before or in combination with other drugs and treatment (such as antibiotics, glucocorticoids, antivirals, oxygen therapy). The IVIG dose and duration of use differed across studies.

**Table 1.** Basic characteristics of the included studies

| Author | Study Location | Study design | Number (M/F) | Disease | Age(range or mean±SD) | Intervention | | Outcome | Risk of bias<sup>#</sup> |
| --- | --- | --- | --- | --- | --- | --- | --- | --- | --- |
|  |  |  |  |  |  | Other treatment<sup>*</sup> | IVIG |  |  |
| Zeng 2003 (33) | Guangzhou | Case series | 5/5 | SARS | 7.3±5.1 | Antibiotics, oxygen inhalation, symptomatic , Comprehensive treatment, etc. | 200-400mg/kg/d for 3 days | I, II, III | 4/8 |
| Wu 2003 (35) | Guangzhou | Case series | 66 | SARS | 16~62 | Antibiotics, glucocorticoids, interferons, antivirals and oxygen therapy. | 5~10 g/d for 3 to 6 days | IV | 4/8 |
| Jann-Tay 2004 (34) | Taiwan | Case series | 22 | SARS | 24~87 | Methylprednisolone | 1g/kg/d for 2 days | V, VI | 5/8 |
| Wu 2005* (36) | Guangzhou | RCT | 15/29 | SARS | Mean:42/43 | Antibiotics, glucocorticoids | 2.5mg/d for 2 days~10mg/d for 13 days | VII, VIII, IX | NA |
| Liu 2020 (31) | Wuhan | Case series | 29/24 | COVID-19 | Mean:55 | Antibiotics, glucocorticoids, interferons, antivirals and oxygen therapy. | NR | X | 5/8 |
| Cao 2020 (32) | Wuhan | Case report | 3 | COVID-19 | 34~56 | NR | 25 g/d for 5 days | XI | NA |

\* Prior to IVIG treatment, the patient received other treatment.
<sup>#</sup> Risk of bias in case series.
Outcome: I: The duration of fever; II: Total peripheral blood WBC ( $10^9/L$ ); III: Time of the lung lesions subsided obviously; IV: Adverse effects; V: WBC counts ( $10^9/L$ ); VI: Platelet counts ( $10^9/L$ ); VII: Serum globulin (g/L); VIII: The incidence of nosocomial infection; IX: The risk of death; X: Survival probability; XI: The progression of disease cascade;
NR: Not Report; NA: Not Applicable.

**Table 2.** GRADE evidence profile

| № of studies | Sample size | Certainty assessment |  |  |  |  | Effect Value<br>(95% CI) | Certainty |
| --- | --- | --- | --- | --- | --- | --- | --- | --- |
|  |  | Risk of bias | Inconsistency | Indirectness | Imprecision | Other considerations |  |  |
| Serum globulin |  |  |  |  |  |  |  |  |
| RCT (31) | 44 | serious <sup>1</sup> | - | serious <sup>2</sup> | - | none | - | ⊕⊕○○<br>LOW |
| Nosocomial infection rate |  |  |  |  |  |  |  |  |
| RCT (31) | 44 | serious <sup>1</sup> | serious <sup>3</sup> | not serious | not serious | none | OR 1.25<br>(0.47 to 3.31) | ⊕⊕○○<br>LOW |
| the risk of death |  |  |  |  |  |  |  |  |
| RCT (31) | 44 | not serious | not serious | not serious | not serious | not serious | - | ⊕⊕○○<br>LOW |

CI: Confidence Interval; OR: Odds Ratio;

**Table 3.**
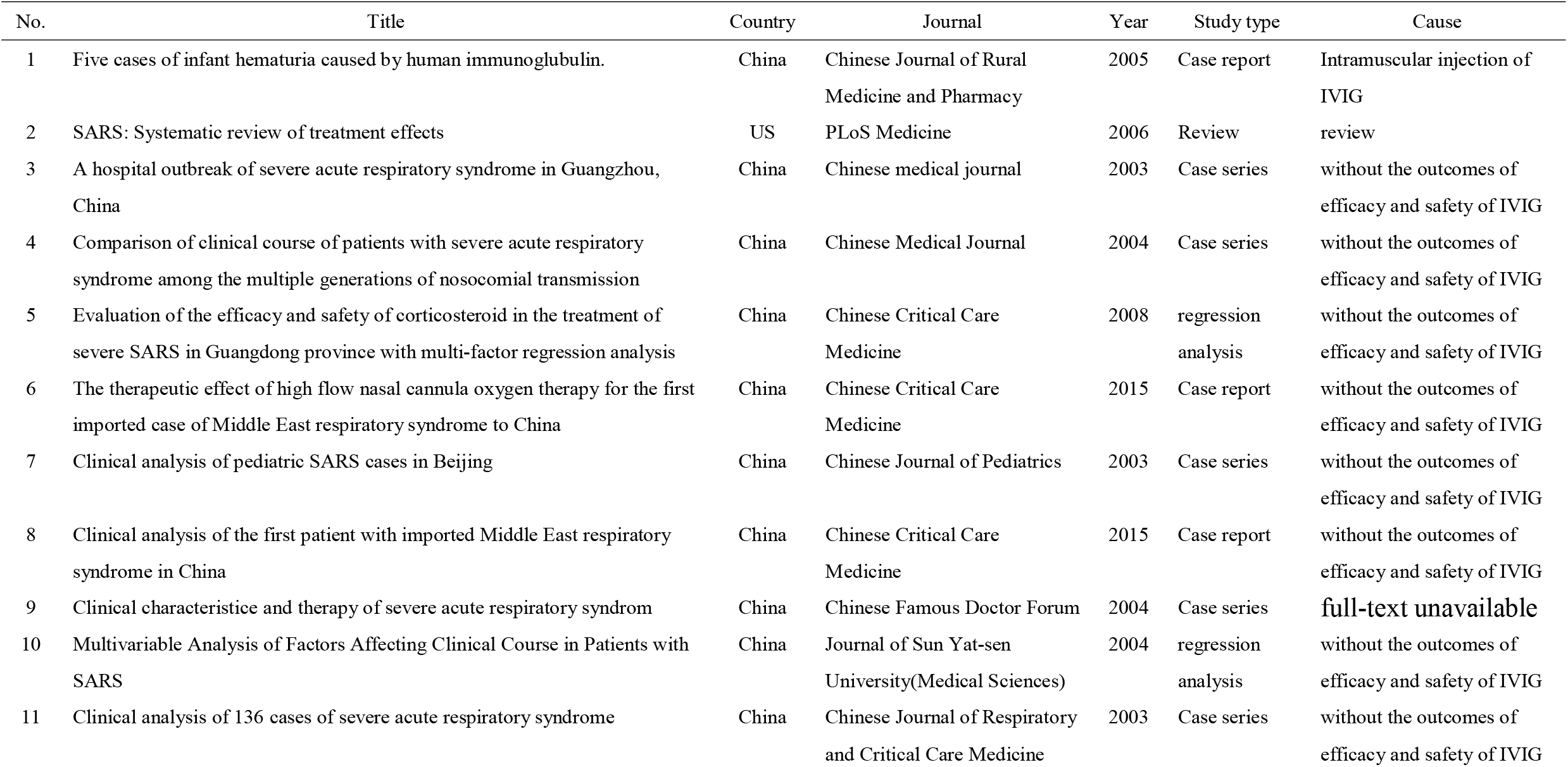
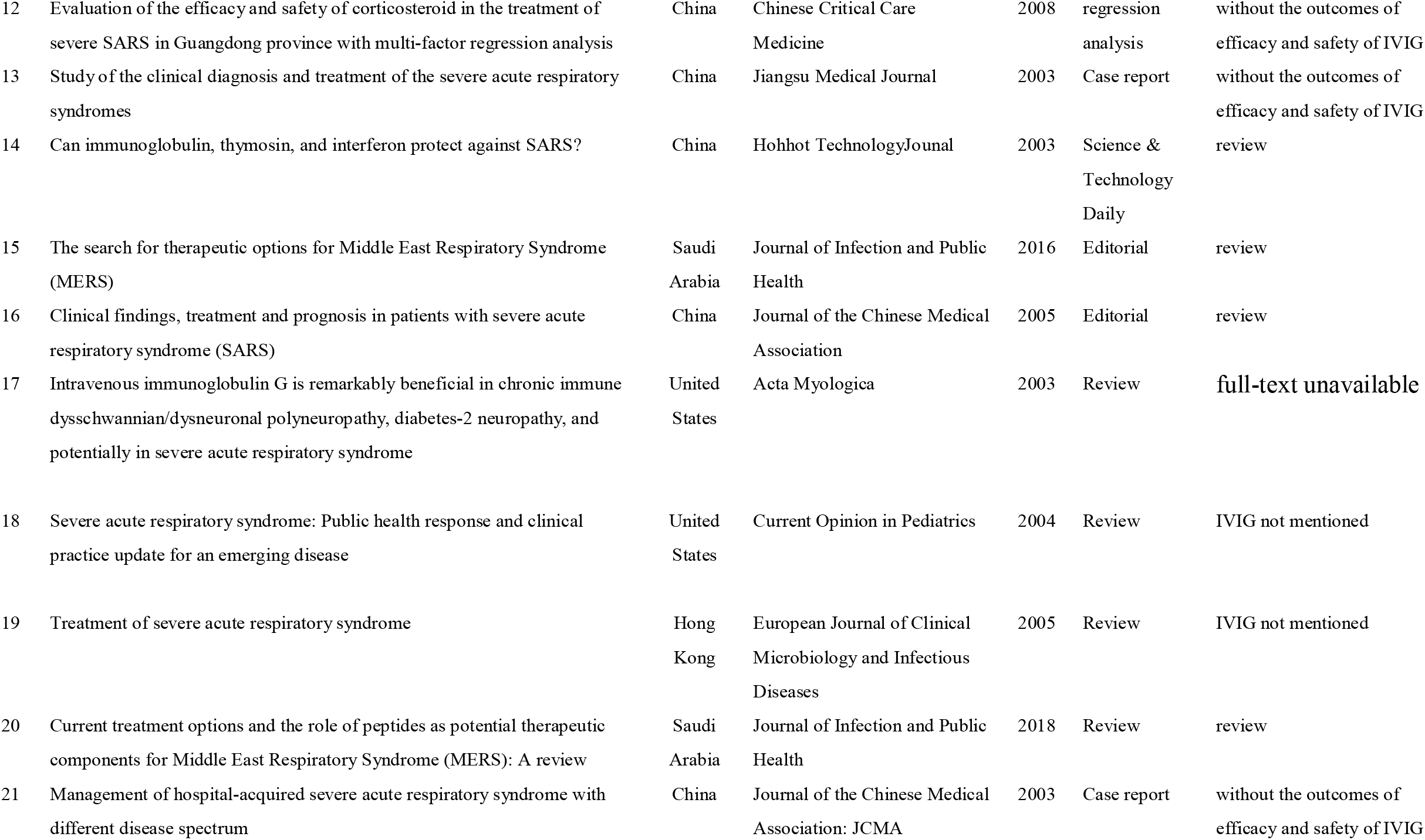
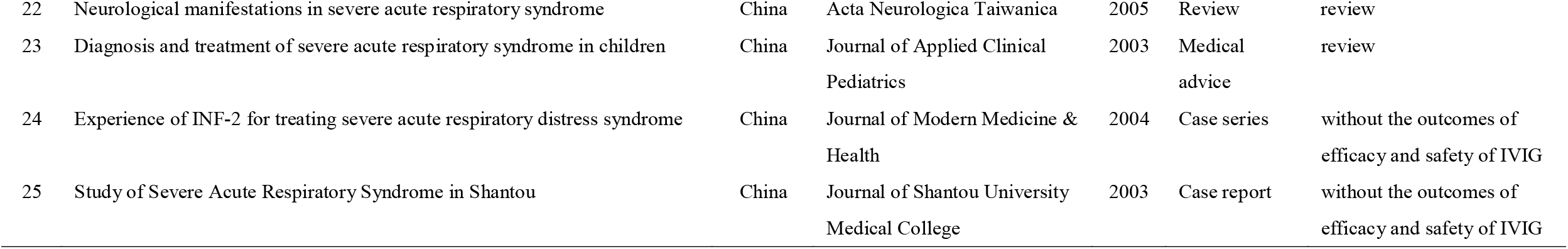
The characteristics of excluded studies

| No. | Title | Country | Journal | Year | Study type | Cause |
| --- | --- | --- | --- | --- | --- | --- |
| 1 | Five cases of infant hematuria caused by human immunoglobulin. | China | Chinese Journal of Rural Medicine and Pharmacy | 2005 | Case report | Intramuscular injection of IVIG |
| 2 | SARS: Systematic review of treatment effects | US | PLoS Medicine | 2006 | Review | review |
| 3 | A hospital outbreak of severe acute respiratory syndrome in Guangzhou, China | China | Chinese medical journal | 2003 | Case series | without the outcomes of efficacy and safety of IVIG |
| 4 | Comparison of clinical course of patients with severe acute respiratory syndrome among the multiple generations of nosocomial transmission | China | Chinese Medical Journal | 2004 | Case series | without the outcomes of efficacy and safety of IVIG |
| 5 | Evaluation of the efficacy and safety of corticosteroid in the treatment of severe SARS in Guangdong province with multi-factor regression analysis | China | Chinese Critical Care Medicine | 2008 | regression analysis | without the outcomes of efficacy and safety of IVIG |
| 6 | The therapeutic effect of high flow nasal cannula oxygen therapy for the first imported case of Middle East respiratory syndrome to China | China | Chinese Critical Care Medicine | 2015 | Case report | without the outcomes of efficacy and safety of IVIG |
| 7 | Clinical analysis of pediatric SARS cases in Beijing | China | Chinese Journal of Pediatrics | 2003 | Case series | without the outcomes of efficacy and safety of IVIG |
| 8 | Clinical analysis of the first patient with imported Middle East respiratory syndrome in China | China | Chinese Critical Care Medicine | 2015 | Case report | without the outcomes of efficacy and safety of IVIG |
| 9 | Clinical characteristic and therapy of severe acute respiratory syndrom | China | Chinese Famous Doctor Forum | 2004 | Case series | **full-text unavailable** |
| 10 | Multivariable Analysis of Factors Affecting Clinical Course in Patients with SARS | China | Journal of Sun Yat-sen University(Medical Sciences) | 2004 | regression analysis | without the outcomes of efficacy and safety of IVIG |
| 11 | Clinical analysis of 136 cases of severe acute respiratory syndrome | China | Chinese Journal of Respiratory and Critical Care Medicine | 2003 | Case series | without the outcomes of efficacy and safety of IVIG |
| 12 | Evaluation of the efficacy and safety of corticosteroid in the treatment of severe SARS in Guangdong province with multi-factor regression analysis | China | Chinese Critical Care Medicine | 2008 | regression analysis | without the outcomes of efficacy and safety of IVIG |
| 13 | Study of the clinical diagnosis and treatment of the severe acute respiratory syndromes | China | Jiangsu Medical Journal | 2003 | Case report | without the outcomes of efficacy and safety of IVIG |
| 14 | Can immunoglobulin, thymosin, and interferon protect against SARS? | China | Hohhot Technology Journal | 2003 | Science & Technology Daily | review |
| 15 | The search for therapeutic options for Middle East Respiratory Syndrome (MERS) | Saudi Arabia | Journal of Infection and Public Health | 2016 | Editorial | review |
| 16 | Clinical findings, treatment and prognosis in patients with severe acute respiratory syndrome (SARS) | China | Journal of the Chinese Medical Association | 2005 | Editorial | review |
| 17 | Intravenous immunoglobulin G is remarkably beneficial in chronic immune dysschwannian/dysneuronal polyneuropathy, diabetes-2 neuropathy, and potentially in severe acute respiratory syndrome | United States | Acta Myologica | 2003 | Review | full-text unavailable |
| 18 | Severe acute respiratory syndrome: Public health response and clinical practice update for an emerging disease | United States | Current Opinion in Pediatrics | 2004 | Review | IVIG not mentioned |
| 19 | Treatment of severe acute respiratory syndrome | Hong Kong | European Journal of Clinical Microbiology and Infectious Diseases | 2005 | Review | IVIG not mentioned |
| 20 | Current treatment options and the role of peptides as potential therapeutic components for Middle East Respiratory Syndrome (MERS): A review | Saudi Arabia | Journal of Infection and Public Health | 2018 | Review | review |
| 21 | Management of hospital-acquired severe acute respiratory syndrome with different disease spectrum | China | Journal of the Chinese Medical Association: JCMA | 2003 | Case report | without the outcomes of efficacy and safety of IVIG |
| 22 | Neurological manifestations in severe acute respiratory syndrome | China | Acta Neurologica Taiwanica | 2005 | Review | review |
| 23 | Diagnosis and treatment of severe acute respiratory syndrome in children | China | Journal of Applied Clinical Pediatrics | 2003 | Medical advice | review |
| 24 | Experience of INF-2 for treating severe acute respiratory distress syndrome | China | Journal of Modern Medicine & Health | 2004 | Case series | without the outcomes of efficacy and safety of IVIG |
| 25 | Study of Severe Acute Respiratory Syndrome in Shantou | China | Journal of Shantou University Medical College | 2003 | Case report | without the outcomes of efficacy and safety of IVIG |

### Risk of bias for included studies

We found a high risk of bias in random sequencing, allocation concealment and blinding in the only included RCT. All case series had a moderate risk (score 4 to 5 out of 8), one case report meeting 8 of the 8 items of the JBI quality appraisal tool (*Table 1*).

### Quality of the evidence

The quality of evidence for all outcomes assessed in the only included RCT was graded low (*Table 2*), primarily due to serious risk of bias and imprecision. As we included four case series, we judged that reporting a ‘GRADE evidence profile’ would not be meaningful. Overall, the quality of evidence was very low for most outcomes and cannot thus provide a reliable indication of any likely effect across outcomes.

### COVID-19

A case series of 109 adults with COVID-19 reported that most patients used antibiotics and antiviral treatment, and over half of the patients were given glucocorticoid therapy and IVIG. The survival probability of patients with ARDS could not be improved by antiviral, glucocorticoid, or immunoglobulin treatment. The risk of death was not associated with the use of IVIG in the patients with ARDS (31).A case report of three adults with COVID-19 reported that a high dose IVIG (25 g/d for 5 days) administered at the appropriate point could successfully block the progression of disease cascade (result of the clinical symptoms, laboratory inspection indicators and chest CT scan), and finally improve the outcome of COVID 19 (32).

### SARS

#### The incidence of nosocomial infection and the risk of death

The randomized controlled trial of 44 adults with severe SARS found no significant difference in the risk of death (18.1% vs. 23.8%) or the incidence of nosocomial infection (65.2% vs. 52.4%) between adults treated either with IVIG or with conventional treatment. And there was no significant difference in the incidence of nosocomial infection between ALI (50.0% vs. 38.5%) and ARDS (81.8% vs. 75.0%) patients (36).

### Laboratory inspection

One case series reported the patients with SARS who did not receive steroids for severe hemocytopenia had increased WBC counts and platelet counts after undergoing IVIG (34). Another case series reported the children with persistent fever who were given

IVIG had significantly improved total peripheral blood WBC after undergoing IVIG (33). The included RCT showed the serum globulin increased slightly in the IVIG group, but decreased in the conventional treatment group, the difference was not significant (36).

### The duration of fever

One case series included ten children with persistent fever who were given IVIG. The body temperature ranged between 38.4°C and 40°C at baseline and the duration of fever was 1 to 4 days after IVIG (33).

## Imaging testing

One case series reported that chest radiographs in children who were given IVIG showed more patchy focal asymmetric infiltrative shadows, more rapid time of the lung lesions subsided obviously than in a randomly selected, age- and sex-matched control group of 20 children without IVIG (33).

### Adverse effects

One case series reported no adverse effects associated with IVIG, when IVIG was used in the patients who had high fever or other obvious poisoning symptoms, who were in the early stage of the disease, who were treated with glucocorticoids, or whose white blood cell counts below 3.0 ×10^9^ /L (35).

## Discussion

We only found limited evidence about the use of IVIG to treat children or adults with severe COVID-19. Since SARS and COVID-19 belong to the same family of viruses, we used IVIG treatment of SARS as indirect evidence, even though the quality of the included studies was generally low. The results were also inconsistent, and no benefit was found in the only identified randomized controlled trial.

An earlier systematic review of treatment effects with SARS concluded that although four studies suggested an improvement in the patients’ condition after IVIG treatment, more controlled trials are needed to provide evidence of the potential benefits on IVIG against SARS. The results of the review are roughly in line with our findings, more high-quality evidence about the benefits and disadvantages of IVIG for COVID-19 and SARS are needed (37).

There was no apparent benefit from IVIG, despite it being used to treat other respiratory infections. A meta-analysis of seven RCTs in children aged less than three years with respiratory syncytial virus infection found no evidence of differences between children treated with immunoglobulin or with placebo in the risk of death (RR=0.87, 95% CI: 0.14,5.27) or serious adverse events (RR=1.08, 95% CI: 0.65,1.79), or in the duration of hospitalization (MD=-0.70, 95% CI: −1.83,0.42) (38). SARS belongs to the category of systemic inflammatory response syndromes (SIRS), and severe SARS often manifests as ALI, ARDS and progresses to severe sepsis (39,40). The course of COVID-19 may also be similar. A meta-analysis of nine RCTs showed that IVIG did not reduce the mortality (OR=0.95, 95% CI: 0.80,1.13), length of hospital stay (MD=-4.08, 95% CI: −6.47,−1.69]), or the risk of death or major disability before two years of age (RR=0.98,95% CI: 0.88,1.09]) in infants with suspected or confirmed infection, compared with placebo or no intervention (41).

IVIG is prepared from pools of plasma obtained from several thousand healthy blood donors. Unlike convalescent plasma from patients with COVID-19, IVIG does not contain SARS-Cov-2 neutralizing antibody (42,43). The review showed that there was no evidence that IVIG has an effect on anti-MERS-CoV, or that IVIG would cause kidney failure or thrombosis in patients with MERS (44). IVIG could increase the risk of vaccination delay. A study by National Advisory Committee on Immunization and the American Advisory Committee on Immunization showed a delay of five months of varicella vaccine in patients who received IVIG and varicella immune globulin (VZIG) (45). IVIG may increase the risk of infections transmitted by transfusion (42). Some adverse effects, such as thrombosis, aseptic meningitis, hemolysis, and renal failure, are mainly associated with the use of high-dose IVIG (46).

The condition, dose and duration of IVIG were inconsistent between studies, the efficacy, and the associated adverse effects remain unclear. The first severe case of COVID-19 in children in China took IVIG with a dose of 400 mg/kg for a duration of five days (8). The recommended dosage of IVIG for children with severe COVID-19 was also inconsistent in different guidelines, including 1.0g/kg/d for 2 days, or 400mg/kg/d for 5 days, 0.2g/kg/d for 3∼5 days, or 1∼2g/kg for 2∼3 days (47-49). Therefore, it is particularly important and urgent to study the benefits and disadvantages of IVIG treatment in children with COVID-19. It is promising that a trial addressing efficacy and safety of IVIG therapy in patients with severe or critical COVID-19 disease has been registered on (50). Randomized, double-blinded, large sample, multicenter clinical trials on children are urgently needed for getting scientific evidence to support clinical decision-making.

### Strength and limitations

This is the first systematic review of IVIG treatment for children with COVID-19. There are several limitations in this systematic review. First, the use of glucocorticoids or a combination of a variety of broad-spectrum antibiotics before IVIG may lead to changes in the microecology of the body, affect the immune regulation function, and thus also affect the effect of IVIG. Second, the total sample size of this study was insufficient to make strong conclusions, and the quality of the methodology was generally low which affect the certainty of the results. Finally, we may have missed some studies as we only included studies published in Chinese and English.

### Conclusion

There is no direct evidence for IVIG in children with COVID-19, current evidence is insufficient to assess the effectiveness and safety of IVIG for children with severe COVID-19. Therefore, we cannot suggest use of IVIG for the treatment of COVID-19 in children. More clinical studies to address this topic are needed.

## Author Contributions

(I) Conception and design: Y Chen, and E Liu; (II) Administrative support: K Yang and Z Fu; (III) Provision of study materials or patients: J Zhang, Y Yang; (IV) Collection and assembly of data: Y Yang and N Yang; (V) Data analysis and interpretation: Y Ma and J Zhang; (VI) Manuscript writing: All authors; (VII) Final approval of manuscript: All authors.

## Data Availability

This study is a rapid review based on the original researches. All the data are from the existing studies and are true.

## Acknowledgments

We thank Janne Estill, Institute of Global Health of University of Geneva for providing guidance and comments for our review. We thank all the authors for their wonderful collaboration.

## Footnote

### Ethical Statement

The authors are accountable for all aspects of the work in ensuring that questions related to the accuracy or integrity of any part of the work are appropriately investigated and resolved.

## Supplementary Material 1-Search strategy

### EMBASE

#1 ‘middle east respiratory syndrome coronavirus’/exp #2 ‘severe acute respiratory syndrome’/exp

#3 ‘sars coronavirus’/exp

#4 ‘COVID-19’:ab,ti

#5 ‘SARS-COV-2’:ab,ti

#6 ‘novel coronavirus’:ab,ti

#7 ‘2019-novel coronavirus’:ab,ti

#8 ‘coronavirus disease-19’:ab,ti

#9 ‘coronavirus disease 2019’:ab,ti

#10 ‘COVID 19’:ab,ti

#11 ‘novel cov’:ab,ti

#12 ‘2019-ncov’:ab,ti

#13 ‘2019-cov’:ab,ti

#14 ‘wuhan-cov’:ab,ti

#15 ‘wuhan coronavirus’:ab,ti

#16 ‘wuhan seafood market pneumonia virus’:ab,ti

#17 ‘middle east respiratory syndrome’:ab,ti

#18 ‘middle east respiratory syndrome coronavirus’:ab,ti

#19 ‘mers’:ab,ti

#20 ‘mers-cov’:ab,ti

#21 ‘severe acute respiratory syndrome’:ab,ti

#22 ‘sars’:ab,ti

#23 ‘sars-cov’:ab,ti

#24 ‘sars-related’:ab,ti

#25 ‘sars-associated’:ab,ti

#26 #1-#25 / OR

#27 ‘Immunoglobulins’/exp

#28 ‘Intravenous Immunoglobulin*’:ab,ti

#29 ‘Intravenous IG’:ab,ti

#30 ‘immune globulin*’:ab,ti

#31 ‘IVIG’:ab,ti

#32 ‘IV Immunoglobulin*’:ab,ti

#33 ‘Intravenous Antibodies’:ab,ti

#34 ‘gamma globulin*’:ab,ti

#35 ‘gamma-globulin*’:ab,ti

#36 ‘Flebogamma DIF’:ab,ti

#37 ‘Gamunex’:ab,ti

#38 ‘Globulin-N’:ab,ti

#39 ‘Globulin N’:ab,ti

#40 ‘Intraglobin’:ab,ti

#41 ‘Gammagard’:ab,ti

#42 ‘Gamimune’:ab,ti

#43 ‘Gamimmune’:ab,ti

#44 ‘Privigen’:ab,ti

#45 ‘Sandoglobulin’:ab,ti

#46 ‘Venoglobulin’:ab,ti

#47 ‘Iveegam’:ab,ti

#48 ‘Endobulin’:ab,ti

#49 ‘Gammonativ’:ab,ti

#50 27-49/OR

#51 #26 AND #50

### PubMed

#1 “COVID-19” [Supplementary Concept]

#2 “Severe Acute Respiratory Syndrome Coronavirus 2” [Supplementary Concept]

#3 “Middle East Respiratory Syndrome Coronavirus” [Mesh]

#4 “Severe Acute Respiratory Syndrome” [Mesh]

#5 “SARS Virus” [Mesh]

#6 “COVID-19” [Title/Abstract]

#7 “SARS-COV-2” [Title/Abstract]

#8 “Novel coronavirus” [Title/Abstract]

#9 “2019-novel coronavirus” [Title/Abstract]

#10 “coronavirus disease-19” [Title/Abstract]

#11 “coronavirus disease 2019” [Title/Abstract]

#12 “COVID 19” [Title/Abstract]

#13 “Novel CoV” [Title/Abstract]

#14 “2019-nCoV” [Title/Abstract]

#15 “2019-CoV” [Title/Abstract]

#16 “Wuhan-Cov” [Title/Abstract]

#17 “Wuhan Coronavirus” [Title/Abstract]

#18 “Wuhan seafood market pneumonia virus” [Title/Abstract]

#19 “Middle East Respiratory Syndrome” [Title/Abstract]

#20 “MERS” [Title/Abstract]

#21 “MERS-CoV” [Title/Abstract]

#22 “Severe Acute Respiratory Syndrome” [Title/Abstract]

#23 “SARS” [Title/Abstract]

#24 “SARS-CoV” [Title/Abstract]

#25 “SARS-Related” [Title/Abstract]

#26 “SARS-Associated” [Title/Abstract]

#27 #1-#26/ OR

#28 “Immunoglobulins, Intravenous” [Mesh]

#29 “gamma-Globulins” [Mesh]

#30 “Intravenous Immunoglobulin*” [Title/Abstract]

#31 “Intravenous IG” [Title/Abstract]

#32 “immune globulin*” [Title/Abstract]

#33 IVIG [Title/Abstract]

#34 “IV Immunoglobulin*” [Title/Abstract]

#35 “Intravenous Antibodies” [Title/Abstract]

#36 “gamma globulin*” [Title/Abstract]

#37 “gamma-globulin*” [Title/Abstract]

#38 “Flebogamma DIF” [Title/Abstract]

#39 Gamunex [Title/Abstract]

#40 “Globulin-N” [Title/Abstract]

#41 “Globulin N” [Title/Abstract]

#42 Intraglobin [Title/Abstract]

#43 Gammagard [Title/Abstract]

#44 Gamimune [Title/Abstract]

#45 Gamimmune [Title/Abstract]

#46 Privigen [Title/Abstract]

#47 Sandoglobulin [Title/Abstract]

#48 Venoglobulin [Title/Abstract]

#49 Iveegam [Title/Abstract]

#50 Endobulin [Title/Abstract]

#51 Gammonativ [Title/Abstract]

#52 28-#51/ OR

#53 #27 AND #52

### Cochrane library

#1 MeSH descriptor: [Middle East Respiratory Syndrome Coronavirus] explode all trees

#2 MeSH descriptor: [Severe Acute Respiratory Syndrome] explode all trees

#3 MeSH descriptor: [SARS Virus] explode all trees

#4 “COVID-19”:ti,ab,kw

#5 “SARS-COV-2”:ti,ab,kw

#6 “Novel coronavirus”:ti,ab,kw

#7 “2019-novel coronavirus”:ti,ab,kw

#8 “Novel CoV”:ti,ab,kw

#9 “2019-nCoV”:ti,ab,kw

#10 “2019-CoV”:ti,ab,kw

#11 “coronavirus disease-19”:ti,ab,kw

#12 “coronavirus disease 2019”:ti,ab,kw

#13 “COVID 19”:ti,ab,kw

#14 “Wuhan-Cov”:ti,ab,kw

#15 “Wuhan Coronavirus”:ti,ab,kw

#16 “Wuhan seafood market pneumonia virus”:ti,ab,kw

#17 “Middle East Respiratory Syndrome”:ti,ab,kw

#18 “MERS”:ti,ab,kw

#19 “MERS-CoV”:ti,ab,kw

#20 “Severe Acute Respiratory Syndrome”:ti,ab,kw

#21 “SARS”:ti,ab,kw

#22 “SARS-CoV”:ti,ab,kw

#23 “SARS-Related”:ti,ab,kw

#24 “SARS-Associated”:ti,ab,kw

#25 #1-#24/ OR

#26 MeSH descriptor: [Immunoglobulins, Intravenous] explode all trees

#27 MeSH descriptor: [gamma-Globulins] explode all trees

#28 “Intravenous Immunoglobulin*” :ti,ab,kw

#29 “Intravenous IG”:ti,ab,kw

#30 “immune globulin*”:ti,ab,kw

#31 “IVIG”:ti,ab,kw

#32 “IV Immunoglobulin*”:ti,ab,kw

#33 “Intravenous Antibodies”:ti,ab,kw

#34 “gamma globulin*”:ti,ab,kw

#35 “gamma-globulin*”:ti,ab,kw

#36 “Flebogamma DIF”:ti,ab,kw

#37 “Gamunex”:ti,ab,kw

#38 “Globulin-N”:ti,ab,kw

#39 “Globulin N”:ti,ab,kw

#40 “Intraglobin”:ti,ab,kw

#41 “Gammagard”:ti,ab,kw

#42 “Gamimune”:ti,ab,kw

#43 “Gamimmune”:ti,ab,kw

#44 “Privigen”:ti,ab,kw

#45 “Sandoglobulin”:ti,ab,kw

#46 “Venoglobulin”:ti,ab,kw

#47 “Iveegam”:ti,ab,kw

#48 “Endobulin”:ti,ab,kw

#49 “Gammonativ”:ti,ab,kw

#50 26-#49/ OR

#51 #25 AND #50

### Web of Science

#1 TOPIC: “COVID-19”

#2 TOPIC: “SARS-COV-2”

#3 TOPIC: “Novel coronavirus”

#4 TOPIC: “2019-novel coronavirus”

#5 TOPIC: “coronavirus disease-19” [Title/Abstract]

#6 TOPIC: “coronavirus disease 2019” [Title/Abstract]

#7 TOPIC: “COVID 19” [Title/Abstract]

#8 TOPIC: “Novel CoV”

#9 TOPIC: “2019-nCoV”

#10 TOPIC: “2019-CoV”

#11 TOPIC: “Wuhan-Cov”

#12 TOPIC: “Wuhan Coronavirus”

#13 TOPIC: “Wuhan seafood market pneumonia virus”

#14 TOPIC: “Middle East Respiratory Syndrome”

#15 TOPIC: “MERS”

#16 TOPIC: “MERS-CoV”

#17 TOPIC: “Severe Acute Respiratory Syndrome”

#18 TOPIC: “SARS”

#19 TOPIC: “SARS-CoV”

#20 TOPIC: “SARS-Related”

#21 TOPIC: “SARS-Associated”

#22 #1-#21 /OR

#23 TOPIC: “Intravenous Immunoglobulin*”

#24 TOPIC: “Intravenous IG”

#25 TOPIC: “immune globulin*”

#26 TOPIC: “IVIG”

#27 TOPIC: “IV Immunoglobulin*”

#28 TOPIC: “Intravenous Antibodies”

#29 TOPIC: “gamma globulin*”

#30 TOPIC: “gamma-globulin*”

#31 TOPIC: “Flebogamma DIF”

#32 TOPIC: “Gamune”

#33 TOPIC: “Globulin-N”

#34 TOPIC: “Globulin N”

#35 TOPIC: “Intraglobin”

#36 TOPIC: “Gammagard”

#37 TOPIC: “Gamimune”

#38 TOPIC: “Gamimmune”

#39 TOPIC: “Privigen”

#40 TOPIC: “Sandoglobulin”

#41 TOPIC: “Venoglobulin”

#42 TOPIC: “Iveegam”

#43 TOPIC: “Endobulin”

#44 TOPIC: “Gammonativ”

#45 #23-#44 /OR

#46 #22 AND #45

### CBM

#1 “新型冠状病毒”[常用字段:智能]

#2 “COVID-19”[常用字段:智能]

#3 “COVID 19”[常用字段:智能]

#4 “2019-nCoV”[常用字段:智能]

#5 “2019-CoV”[常用字段:智能]

#6 “SARS-CoV-2”[常用字段:智能]

#7 “武汉冠状病毒”[常用字段:智能]

#8 “中东呼吸综合征冠状病毒”[不加权:扩展]

#9 “中东呼吸综合征”[常用字段:智能]

#10 “MERS”[常用字段:智能]

#11 “MERS-CoV”[常用字段:智能]

#12 “严重急性呼吸综合征”[不加权:扩展]

#13 “SARS 病毒”[不加权:扩展]

#14 “严重急性呼吸综合征”[常用字段:智能]

#15 “SARS”[常用字段:智能]

#16 #1-#15/ OR

#17 丙种球蛋白[常用字段:智能]

#18 静脉丙球[常用字段:智能]

#19 免疫球蛋白[常用字段:智能]

#20 IVIG[常用字段:智能]

#21 “免疫球蛋白类”[不加权:扩展]

#22 #17-#21/ OR

#23 #16 AND #22

万方

#1 新型冠状病毒[主题]

#2 COVID-19[主题]

#3 COVID 19[主题]

#4 2019-nCoV[主题]

#5 2019-CoV[主题]

#6 SARS-CoV-2[主题]

#7 武汉冠状病毒[主题]

#8 中东呼吸综合征[主题]

#9 MERS[主题]

#10 MERS-CoV[主题]

#11 严重急性呼吸综合征[主题]

#12 SARS[主题]

#13 #1-#12/ OR

#14 丙种球蛋白[主题]

#15 静脉丙球[主题]

#16 IVIG[主题]

#17 免疫球蛋白[主题]

#18 #14-#17/OR

#19 #13 AND #18

### CNKI

#1 “新型冠状病毒”[主题]

#2 “COVID-19”[主题]

#3 “COVID 19”[主题]

#4 “2019-nCoV”[主题]

#5 “2019-CoV”[主题]

#6 “SARS-CoV-2”[主题]

#7 “武汉冠状病毒”[主题]

#8 “中东呼吸综合征”[主题]

#9 “MERS”[主题]

#10 “MERS-CoV”[主题]

#11 “严重急性呼吸综合征”[主题]

#12 “SARS”[主题]

#13 #1-#12/ OR

#14 “丙种球蛋白”[主题]

#15 “静脉丙球”[主题]

#16 “免疫球蛋白”[主题]

#17 “IVIG”[主题]

#18 #13-#17/ OR

#19 #13 AND #18

## Supplementary Material 2

### Explanations

1. Unclear risk of bias in allocation concealment, random sequence generation and blinding.
2. Using other drugs (such as interferon, hormone, etc.) before intervention
3. The value of square I above 50%.

